# Pamrevlumab did not meet its primary endpoint for ambulatory patients with Duchenne Muscular Dystrophy: the LELANTOS-2 trial

**DOI:** 10.1101/2025.02.17.25321425

**Authors:** Brenda L. Wong, Han C. Phan, Yann Péréon, Silvana De Lucia, Yi Dai, Ziyi Chen, Craig Campbell, Valeria A. Sansone, Stéphanie Delstanche, Steven Wang, Olga V. Gambetti, John F. Brandsema, Ewa Carrier, LELANTOS-2 study investigators

**Affiliations:** University of Massachusetts Chan Medical School, Worcester, MA, USA; Rare Disease Research, LLC, Atlanta, GA, USA; Centre de Référence Maladies Neuromusculaires AOC, Filnemus, Hôpital Hôtel-Dieu, Nantes, France; Institute I-Motion, Hôpital Armand Trousseau, Paris, France; Department of Neurology, Peking Union Medical College Hospital, Chinese Academy of Medical Sciences, Beijing, China; The First Affiliated Hospital, Sun Yat-sen University, Guangzhou, Guangdong, China; Pediatrics, Epidemiology and Clinical Neurological Sciences, University of Western Ontario, Department of Paediatrics, Children’s Hospital London Health Sciences Centre, London, Ontario, Canada; The NEMO Center in Milan, Neurorehabilitation Unit, ASST Niguarda Hospital, University of Milan, Milan, Italy; University Department of Neurology, Centre Hospitalier Régional de la Citadelle, Liège, Belgium; FibroGen, Inc., San Francisco, CA, USA; The Children’s Hospital of Philadelphia, Perelman School of Medicine, University of Pennsylvania, Philadelphia, PA, USA

**Keywords:** ambulatory, Duchenne muscular dystrophy, North Star Ambulatory Assessment, pamrevlumab

## Abstract

**Background:** In Duchenne muscular dystrophy (DMD), fibrosis is linked to connective tissue growth factor (CTGF) overexpression. Pamrevlumab, a fully human monoclonal antibody that inhibits CTGF activity, showed promise as a DMD treatment in a phase 2 trial.

**Objective:** LELANTOS-2 (NCT04632940) was a global phase 3 study of the safety and efficacy of pamrevlumab for ambulatory males 6 to <12 years old with DMD.

**Methods:** Patients were randomized 1:1 to pamrevlumab 35 mg/kg every 2 weeks for 52 weeks or placebo. All received a stable corticosteroid regimen (deflazacort or prednisone/prednisolone). Primary endpoint was change in North Star Ambulatory Assessment (NSAA) total score from baseline to Week 52. Treatment-emergent adverse events (TEAEs) were noted. Patients who completed the main study period were eligible to enroll in the open-label extension (OLE).

**Results:** Seventy-three patients enrolled (mean [SD] age, 9.0 [1.5] years; pamrevlumab [n=37], placebo [n=36]). Between-group baseline characteristics were similar. The difference in change in NSAA score was not significant (least squares [LS] mean [SE]: pamrevlumab, –3.022 [0.5505] vs placebo, –2.494 [0.6962]; *p*=0.5553). Across all secondary endpoints, there were no significant differences between patients treated with pamrevlumab or placebo. Nearly all patients (pamrevlumab, n=35 [97.2%]; placebo, n=35 [97.2%]) experienced TEAEs (most mild/moderate). Sixty-eight (34 from each original treatment group) patients enrolled in and received pamrevlumab during the OLE. OLE efficacy and safety were consistent with the main study period. No deaths occurred.

**Conclusions:** Pamrevlumab failed to meet its primary endpoint. Its future as a DMD treatment is uncertain.

## INTRODUCTION

In Duchenne muscular dystrophy (DMD), the absence of or deficiency in dystrophin protein leads to progressive muscle deterioration [1–3]. Progressive muscle fibrosis is a key pathophysiologic component of disease progression [4, 5]. Early symptoms of DMD are often noticed between 1 and 3 years of age and include delayed walking, frequent falls, and difficulty running and climbing stairs. Calf and thigh muscles are noticeably bulkier than normal [4, 6, 7]. Ambulation steadily decreases, with most individuals becoming wheelchair dependent by early adolescence. Upper body function declines through adolescence and young adulthood, resulting in loss of independence [2]. Mortality in DMD is usually related to complications associated with cardiomyopathy or respiratory compromise [4, 8, 9]. Life expectancy for individuals with DMD is typically limited to late 20s or early 30s [7], and there are no curative treatments for DMD. Corticosteroids are currently the standard of care and have been shown to delay loss of ambulation by up to three years and improve respiratory function [1, 9–15].

Pamrevlumab is a fully human monoclonal antibody that binds to and inhibits the activity of connective tissue growth factor (CTGF), which is believed to play a substantial role in the fibrosis that affects muscle tissues in DMD [16–20]. In the open-label phase 2 MISSION study of patients with DMD, pamrevlumab demonstrated slower disease progression than expected based on natural disease history and was well tolerated [21]. The results of the phase 2 study supported the initiation of this phase 3 study of the safety and efficacy of pamrevlumab in combination with systemic corticosteroids for ambulatory males with DMD.

The objective of the phase 3 LELANTOS-2 trial (NCT04632940) was to evaluate the safety and efficacy of pamrevlumab for the treatment of ambulatory males with DMD. A companion trial, LELANTOS-1 (NCT04371666) [22], was designed to evaluate pamrevlumab for non-ambulatory males with DMD.

## METHODS

### Study design and patients

LELANTOS-2 was a phase 3, randomized, double-blind, placebo-controlled, multicenter, multinational study that included ambulatory males with DMD aged 6 to <12 years. Patients were randomized 1:1 to receive pamrevlumab IV 35 mg/kg or placebo every 2 weeks for 52 weeks (**Figure 1**). All patients were required to be on a stable dose of systemic corticosteroids. Patients who completed the treatment period were eligible to enroll in an open-label extension (OLE) period, in which they received pamrevlumab 35 mg/kg every 2 weeks until the last patient from the phase 3 study completed the 52-week treatment period or until the OLE was terminated based on sponsor decision or commercial availability of pamrevlumab.

**Figure 1.**
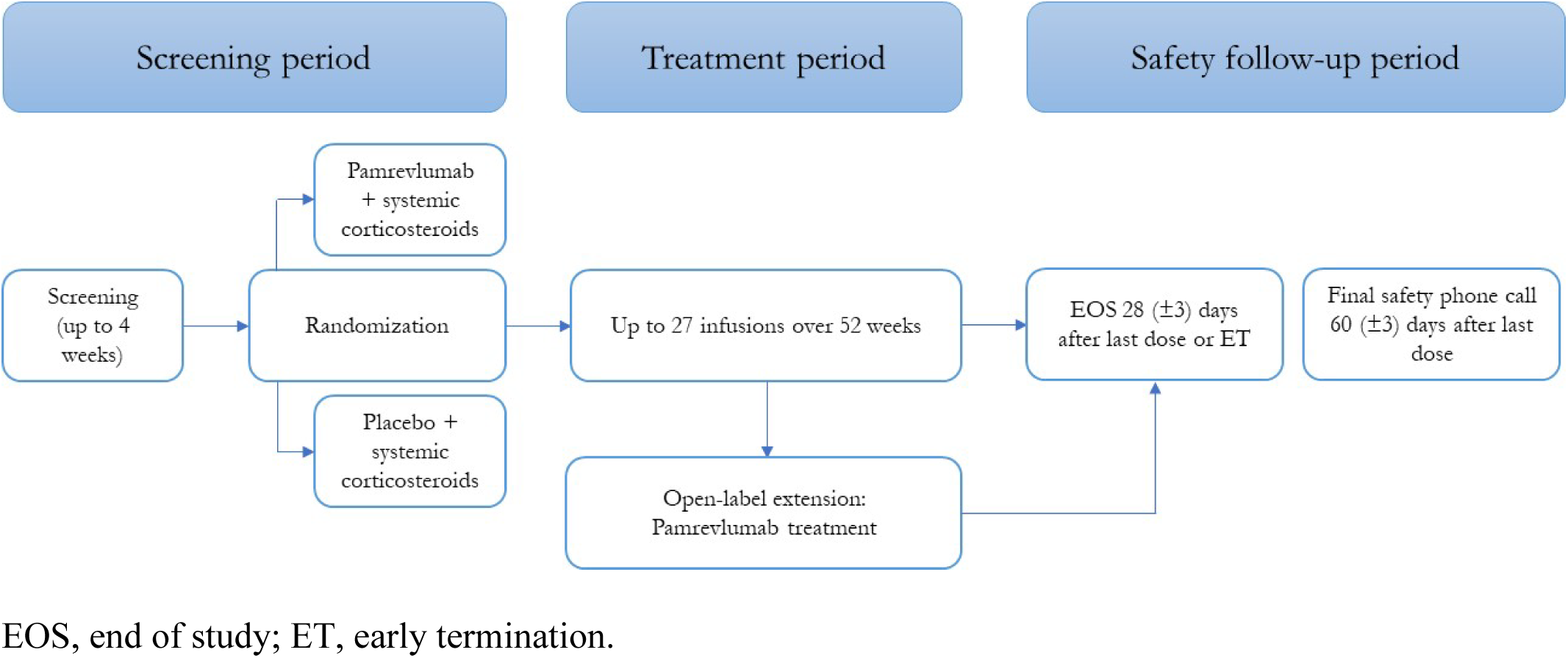
Study design.

### Patients

Males with DMD were included in the study if they were 6 to 12 years old and ambulatory at the time of screening. Full inclusion/exclusion criteria are listed in the **Supplement**.

The intent-to-treat (ITT) set is defined as all randomized patients. Analysis of all efficacy endpoints was based on the ITT set unless otherwise specified. The per-protocol (PP) set is defined as all randomized patients who received at least 24 doses of treatment who had a baseline and at least one post-baseline North Star Ambulatory Assessment (NSAA) assessment, did not terminate the study early, and had no significant protocol deviations. The safety analysis set (SAF) is defined as any patient who received any dose of study medication, and the immunogenicity analysis set (IGS) contains all patients in the SAF with at least one pre-treatment and one post-treatment anti-drug antibody and/or neutralizing antibody (ADA/NAb) assessment.

### Endpoints

The primary efficacy endpoint was change in total score of NSAA from baseline to Week 52. Secondary endpoints included changes from baseline to Week 52 in 4-stair climb velocity (4SCV), 10-meter walk/run test, and time to stand (TTSTAND); time to loss of ambulation from baseline to Week 52; and proportions of patients with >10 sec and ≤10 sec results of 10-meter walk/run test at Week 52. Exploratory endpoints included changes from baseline to Week 52 in Duchenne Video Assessment severity percentage, percent-predicted forced vital capacity and percent-predicted peak expiratory flow, and lower extremities vastus lateralis muscle fibrosis score assessed by magnetic resonance imaging.

Safety endpoints included all treatment-emergent adverse events (TEAEs), treatment-emergent serious adverse events (TESAEs), clinically significant laboratory abnormalities, discontinuation of treatment due to TEAEs, hypersensitivity/anaphylactic reactions, and infusion reactions. Adverse events (AEs) were coded using MedDRA version 26.0 or higher. The number (percentage) of patients with hospitalizations due to any serious adverse event (SAE) with a pulmonary or cardiac cause and the number (percentage) of patients with bone fractures were noted.

### Statistical analysis

All analyses were performed using SAS version 9.4 (Cary, NC, USA) or higher. Categorical variables are presented as number (percentage) with 95% confidence intervals (CIs). Continuous variables are presented as mean (standard deviation [SD] or standard error [SE]) or median (minimum, maximum, or interquartile range) with two-sided 95% CIs.

### Ethics

Informed consent was obtained from each patient or their legal guardian, and the study was approved by the respective Institutional Review Boards of each participating study site (**Table S1**). This study was conducted in accordance with the Declaration of Helsinki, Good Clinical Practice (GCP), the International Council for Harmonisation (ICH) E6 Guidance for GCP, and any other applicable local health and regulatory requirements.

## RESULTS

### Patient disposition

Eighty-eight patients were screened for inclusion, 15 of whom failed screening (**Figure 2**). A total of 73 patients (pamrevlumab, n=37; placebo, n=36) from 37 study sites were enrolled and randomized. Thirty-six patients in each group were treated. In the pamrevlumab group, 35 patients (94.6%) completed 52 weeks of study; two (5.4%) discontinued the study early, both because of withdrawal of consent by patient/guardian. All 36 patients (100%) in the placebo group completed 52 weeks of study. Treatment compliance was high in both groups (median [range]: pamrevlumab, 96.4% [78.7–100.3%]; placebo, 97.8% [74.6–100.3%]). Thirty-four patients (91.9%) in the pamrevlumab group and 35 (97.2%) in the placebo group enrolled in the OLE. Sixty-eight patients (34 from each original treatment group) received pamrevlumab during the OLE.

**Figure 2.**
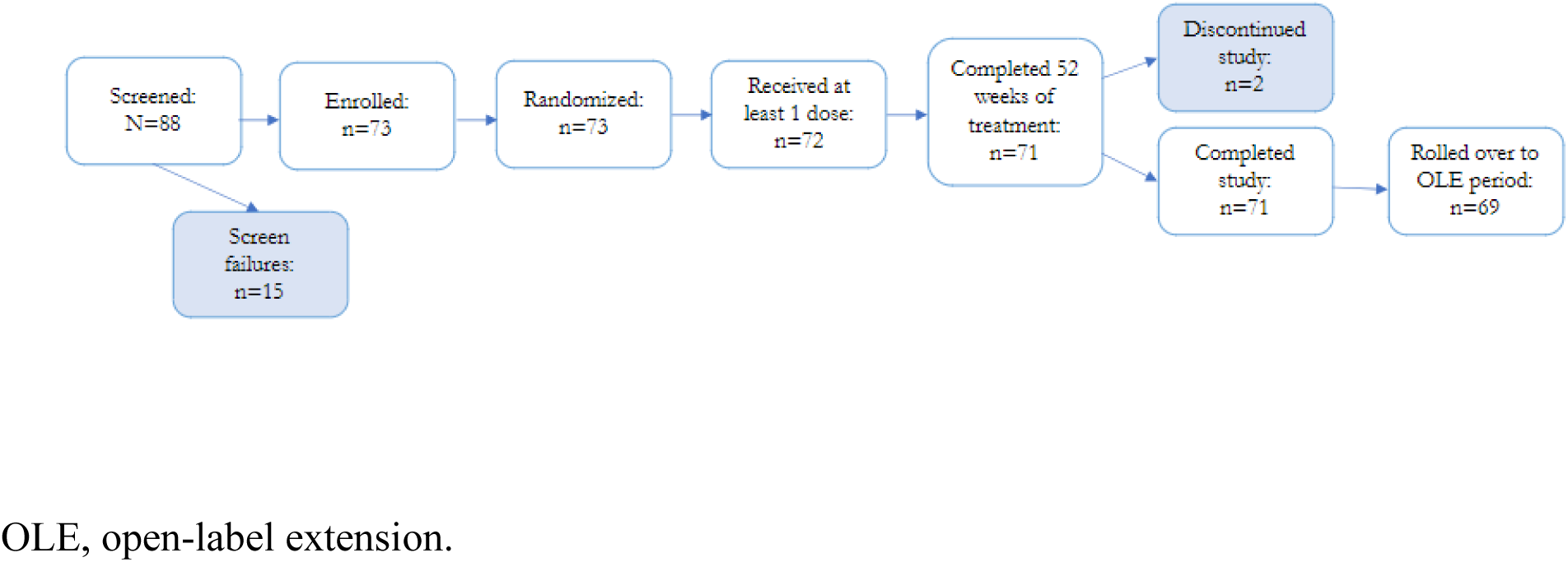
Patient disposition.

The ITT set included 73 patients (pamrevlumab, n=37; placebo, n=36). The PP set included 64 patients (pamrevlumab, n=31; placebo, n=33). The SAF set included 72 patients (pamrevlumab, n=36; placebo, n=36). The IGS set included 12 patients (pamrevlumab, n=5; placebo, n=7). Baseline characteristics were evenly balanced between the pamrevlumab and placebo groups (**Table 1**). The mean (SD) age was 9.0 (1.50) years and most patients (n=45; 61.6%) were White.

**Table 1.** Demographics and disease characteristics.

| *Demographics and baseline clinical characteristics* |  |  |  |  |
| --- | --- | --- | --- | --- |
| Variable |  | Pamrevlumab<br>(n=37) | Placebo<br>(n=36) | Overall<br>(N=73) |
| Age, years | Mean (SD) | 9.1 (1.46) | 9.0 (1.56) | 9.0 (1.50) |
|  | Median (range) | 9.0 (7–12) | 9.0 (6–12) | 9.0 (6–12) |
| Age group, n (%) | ≤9 years old | 22 (59.5) | 22 (61.1) | 44 (60.3) |
|  | >9 years old | 15 (40.5) | 14 (38.9) | 29 (39.7) |
| Sex, n (%) | Male | 37 (100.0) | 36 (100.0) | 73 (100.0) |
| Ethnicity, n (%) | Hispanic or Latino | 1 (2.7) | 5 (13.9) | 6 (8.2) |
|  | Not Hispanic or Latino | 34 (91.9) | 28 (77.8) | 62 (84.9) |
|  | Not reported | 2 (5.4) | 3 (8.3) | 5 (6.8) |
| Race, n (%) | American Indian or Alaska Native | 0 | 0 | 0 |
|  | Asian | 10 (27.0) | 9 (25.0) | 19 (26.0) |
|  | Black or African American | 1 (2.7) | 1 (2.8) | 2 (2.7) |
|  | Native Hawaiian or other Pacific Islander | 0 | 0 | 0 |
|  | White | 23 (62.2) | 22 (61.1) | 45 (61.6) |
|  | Not reported | 2 (5.4) | 2 (5.6) | 4 (5.5) |
|  | Other | 1 (2.7) | 2 (5.6) | 3 (4.1) |
| BMI, kg/m<sup>2</sup> | Mean (SD) | 17.53 (3.088) | 19.45 (3.928) | 18.48 (3.632) |
|  | Median (range) | 16.72 (14.0–25.9) | 18.24 (14.3–30.6) | 17.42 (14.0–30.6) |
| *Disease history and characteristics* |  |  |  |  |
| Age at DMD diagnosis, years | Mean (SD) | 3.7 (1.96) | 4.7 (2.05) | 4.2 (2.05) |
|  | Median (range) | 3.0 (0–9) | 5.0 (1–8) | 4.0 (0–9) |

| ***Demographics and baseline clinical characteristics*** |  |  |  |  |
| --- | --- | --- | --- | --- |
| **Variable** |  | **Pamrevlumab**<br>**(n=37)** | **Placebo**<br>**(n=36)** | **Overall**<br>**(N=73)** |
| Corticosteroid use, n (%) | Prednisone/prednisolone | 12 (32.4) | 18 (50.0) | 30 (41.1) |
|  | Deflazacort | 25 (67.6) | 18 (50.0) | 43 (58.9) |
|  | Daily regimen | 33 (89.2) | 34 (94.4) | 67 (91.8) |
|  | Other regimen | 4 (10.8) | 2 (5.6) | 6 (8.2) |
| Age at first use of corticosteroids, years | Mean (SD) | 4.76 (1.627) | 5.15 (1.398) | 4.95 (1.521) |
|  | Median (range) | 5.00 (1.5–8) | 5.00 (3–7) | 5.00 (1.5–8) |
| Years since first use of corticosteroids | Mean (SD) | 4.32 (2.106) | 3.82 (2.084) | 4.08 (2.096) |
|  | Median (range) | 4.00 (1–10) | 4.00 (1–8) | 4.00 (1–10) |
| Total NSAA score | Mean (SD) | 25.5 (5.04) | 23.6 (6.21) | 24.6 (5.69) |
|  | Median (range) | 26.0 (12–34) | 24.0 (12–34) | 26.0 (12–34) |
| Baseline 10-meter walk/run test, n (%) | ≤5 sec | 20 (54.1) | 18 (50.0) | 38 (52.1) |
|  | >5 sec | 17 (45.9) | 18 (50.0) | 35 (47.9) |
| Baseline time to rise from floor, n (%) | ≤5 sec | 23 (62.2) | 17 (47.2) | 40 (54.8) |
|  | >5 sec | 14 (37.8) | 19 (52.8) | 33 (45.2) |

BMI, body mass index; DMD, Duchenne muscular dystrophy; NSAA, North Star Ambulatory Assessment; SD, standard deviation.

### Efficacy

The least squares (LS) mean (SE) change in total NSAA score for the ITT set was –3.022 (0.5505) for pamrevlumab versus –2.492 (0.6962) for placebo (LS mean difference [SE]: –0.528 [0.8912]; *p*=0.5553) (**Figure 3**). The primary endpoint was not met. Sensitivity, subgroup, and exploratory analyses demonstrated similar results.

**Figure 3.**
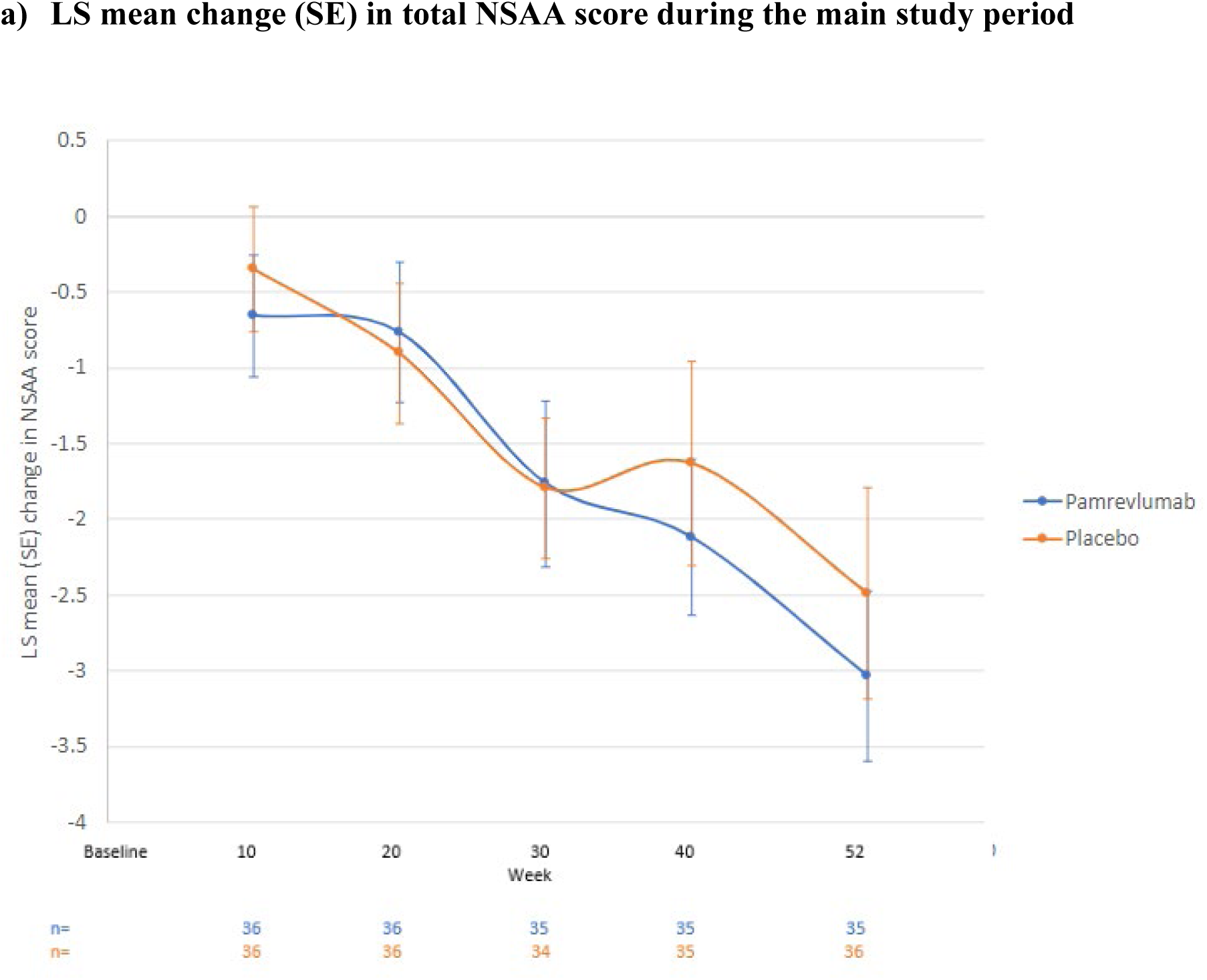
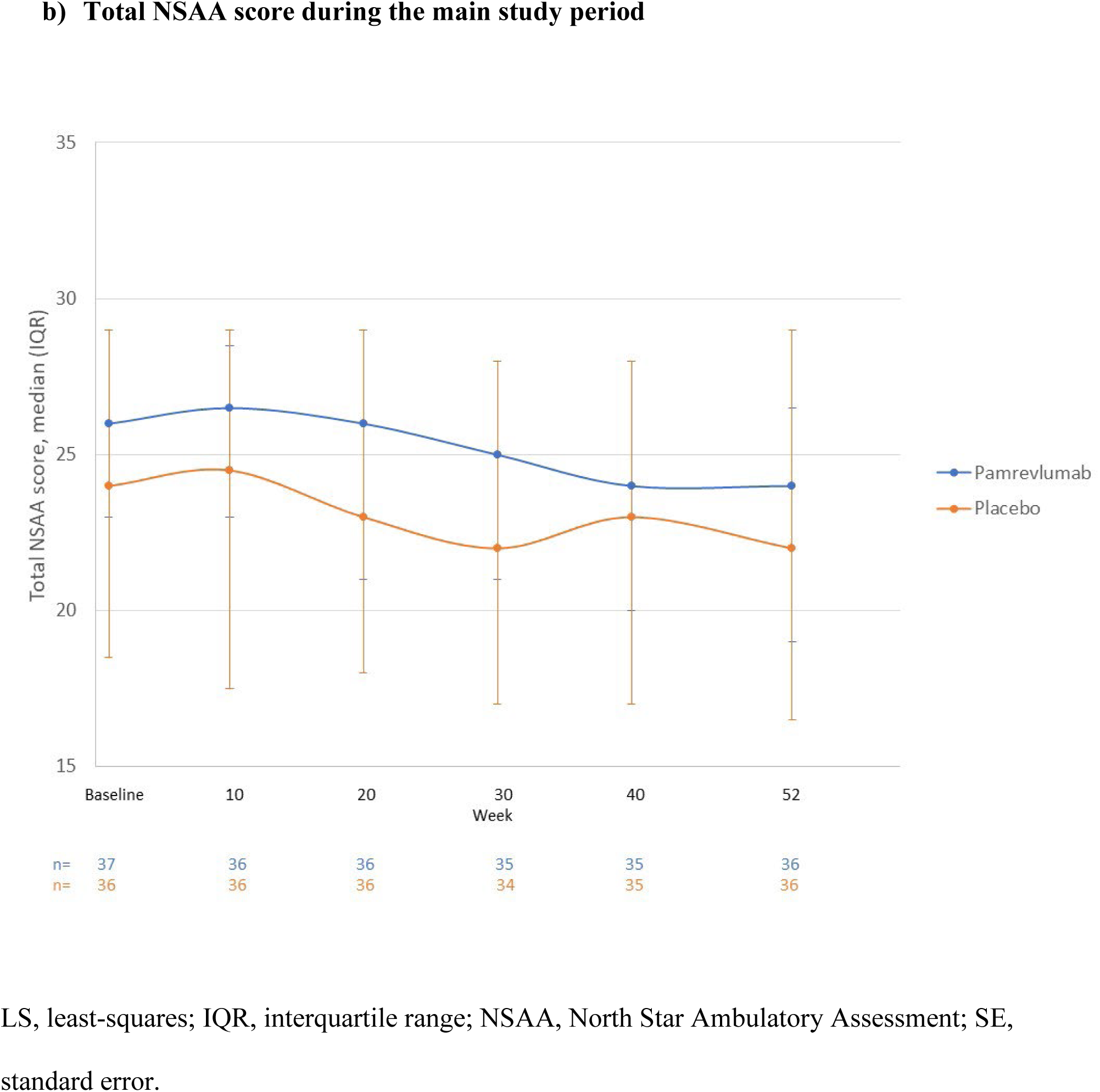
Total NSAA score during the main study period.

None of the secondary or exploratory endpoints demonstrated statistically significant differences (**Table 2**). No patients lost ambulation during the study. At Week 52, one patient (2.8%) in the pamrevlumab group had 10-m walk test >10 sec versus seven (19.4%) in the placebo group (*p*=0.0553). Of note, for the primary endpoint, among patients with a baseline TTSTAND ≤5 seconds (pamrevlumab, n=22; placebo, n=17), NSAA total score decreased over five times more in the pamrevlumab group than in the placebo group (LS mean change [SE], pamrevlumab, – 2.629 [0.8195]; placebo, –0.466 [0.6461]; *p*=0.0435).

**Table 2.** Primary and secondary efficacy endpoints in main study period and OLE.

| **Parameter** | **Change from baseline at Week 52,**<br>**LS mean (SE)** |  |  | **Change from baseline at OLE**<br>**Week 36, LS mean (SE)** |  |  |
| --- | --- | --- | --- | --- | --- | --- |
|  | **Pamrevlumab**<br>**(n=37)** | **Placebo**<br>**(n=36)** | ***p*-**<br>**value** | **Pamrevlumab**<br>**(n=34)** | **Placebo**<br>**(n=34)** | ***p*-**<br>**value** |
| NSAA, total score | *n*=36<br>−3.022 (0.5505) | *n*=36<br>−2.494<br>(0.6962) | 0.5553 | *n*=15<br>−4.457 (0.7602) | *n*=15<br>−4.663<br>(0.8027) | 0.8532 |
| 4-stair climb velocity, cm/sec | *n*=36<br>−2.233 (0.7285) | *n*=36<br>−3.849<br>(0.8645) | 0.1575 | *n*=16<br>−3.410 (1.0816) | *n*=15<br>−5.546<br>(1.1463) | 0.1763 |
| 10-meter walk/run test, m/sec | *n*=36<br>−0.169 (0.0377) | *n*=36<br>−0.197<br>(0.0580) | 0.6862 | *n*=15<br>−0.322 (0.0761) | *n*=14<br>−0.277<br>(0.0686) | 0.6612 |
| TTSTAND, sec | *n*=36<br>2.416 (0.5747) | *n*=31<br>2.743<br>(0.7879) | 0.7391 | *n*=15<br>3.806 (0.6654) | *n*=10<br>3.795<br>(0.8877) | 0.9922 |

LS, least-squares; NSAA, North Star Ambulatory Assessment; OLE, open-label extension; SE, standard error; TTSTAND, time to stand.

OLE efficacy results were consistent with results from the main study, and no strong trends were detected (**Table 2**).

### Safety

The incidence of TEAEs was the same for each treatment group; 35 patients (97.2%) in both the pamrevlumab and placebo groups had at least one TEAE during the study period (**Table 3**). The most common TEAEs for the pamrevlumab treatment group with a ≥5% higher incidence than the placebo group were COVID-19 infection (44.4% vs 22.2%), headache (38.9% vs 13.9%), nasopharyngitis (27.8% vs 19.4%), vomiting (27.8% vs 19.4%), pyrexia (25.0% vs 19.4%), diarrhea (22.2% vs 8.3%), cough (16.7% vs 11.1%), falls (19.4% vs 5.6%), abdominal pain (13.9% vs 8.3%), arthralgia (11.1% vs 5.6%), back pain (11.1% vs 5.6%), and pain in extremity (11.1% vs 5.6%). Four patients (11.1%) in the pamrevlumab group and two (5.6%) in the placebo group experienced Grade ≥3 TEAEs (**Table 3**). The most common TEAEs were infections and infestations for both treatment groups, followed by gastrointestinal disorders, musculoskeletal and connective tissue disorders, and nervous system disorders (**Table 4**). Fourteen patients (38.9%) in the pamrevlumab group and seven (19.4%) in the placebo group had TEAEs considered related to study drug (**Table 3**). The most common treatment-related TEAEs in both groups were gastrointestinal disorders. Three patients (8.3%) in the pamrevlumab group and one (2.8%) in the placebo group experienced TESAEs (**Table 3**). There were no trends in hematology, chemistry, physical examinations, or vital signs results during the study. Four patients (11.1%) in each group experienced bone fractures.

**Table 3.** Safety summary.

| Category | Main study period |  | OLE period |  |
| --- | --- | --- | --- | --- |
|  | Pamrevlumab<br>(n=36), n (%) | Placebo<br>(n=36), n (%) | Pamrevlumab<br>(n=34), n (%) | Placebo<br>(n=34), n (%) |
| Any TEAE | 35 (97.2) | 35 (97.2) | 26 (76.5) | 26 (76.5) |
| Any TEAE leading to discontinuation of study medication | 0 | 0 | 1 (2.9) | 0 |
| Any TEAE leading to interruption of study medication | 15 (41.7) | 8 (22.2) | 2 (5.9) | 4 (11.8) |
| Any TEAE related to study medication (as determined by the investigator) | 14 (38.9) | 7 (19.4) | 7 (20.6) | 3 (8.8) |
| TEAE by maximum severity |  |  |  |  |
| Grade 1/mild | 12 (33.3) | 17 (47.2) | 9 (26.5) | 10 (29.4) |
| Grade 2/moderate | 19 (52.8) | 16 (44.4) | 16 (47.1) | 14 (41.2) |
| Grade 3/severe | 4 (11.1) | 2 (5.6) | 1 (2.9) | 2 (5.9) |
| Grade 4/life threatening | 0 | 0 | 0 | 0 |
| Grade 5/death | 0 | 0 | 0 | 0 |
| Any TESAE | 3 (8.3) | 1 (2.8) | 0 | 0 |
| Any TESAE related to study medication (as determined by the investigator) | 0 | 0 | 0 | 0 |
| Any TESAE leading to death | 0 | 0 | 0 | 0 |
| All reported deaths | 0 | 0 | 0 | 0 |

OLE, open-label extension; TEAE, treatment-emergent adverse event; TESAE, treatment-emergent serious adverse event.

**Table 4.**
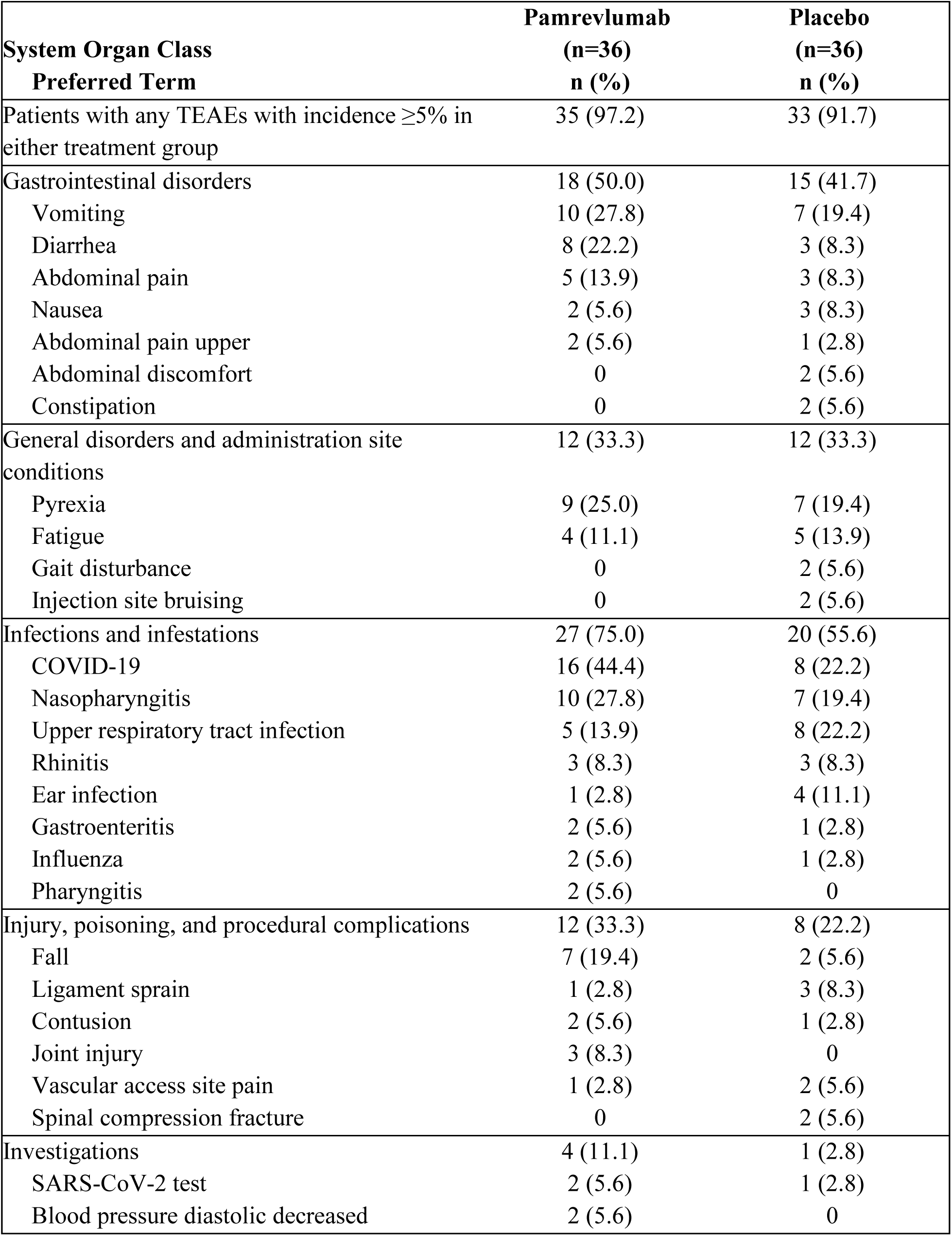
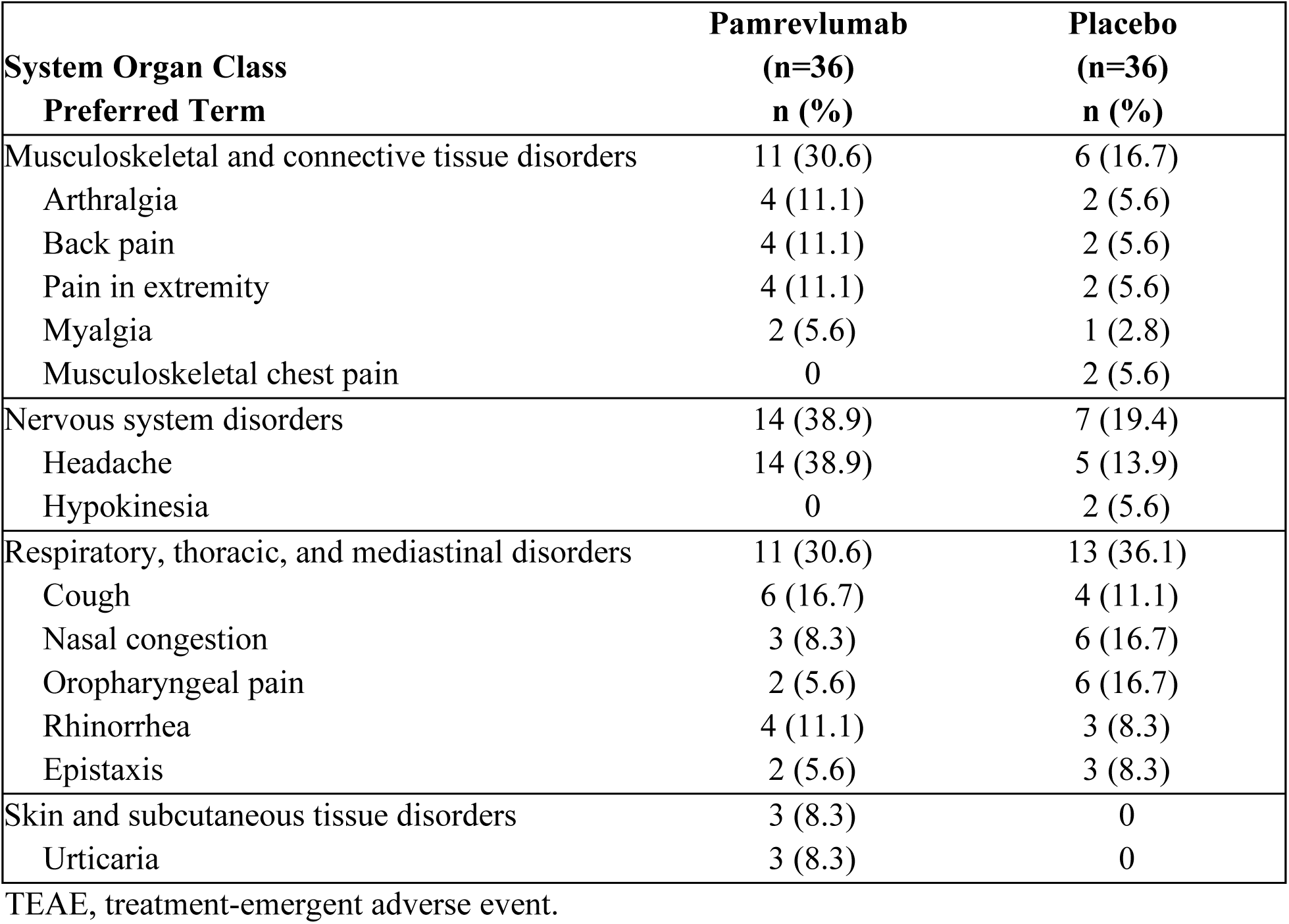
TEAEs with an incidence of ≥5% in either treatment group.

No notable safety trends were observed during the OLE period (**Table 3**). TEAE incidence in the OLE period was equal with the main study treatment groups (pamrevlumab: 76.5%; placebo: 76.5%). In the OLE period, one patient (2.9%) in the pamrevlumab treatment group and two patients (5.9%) in the placebo group had Grade 3 TEAEs. There were no Grade 4 or Grade 5 TEAEs. There were no trends in hematology, chemistry, physical examinations, vital signs, or bone fractures, and no deaths occurred during the OLE period.

Overall, the safety data were in line with expectations and did not reveal any new safety concerns. Pamrevlumab was generally safe and well tolerated.

## DISCUSSION

LELANTOS-2 did not meet its primary endpoint of change in NSAA for ambulatory patients with DMD during the main study, and this trend continued through the OLE. Similarly, none of the secondary endpoints nor the exploratory endpoints demonstrated statistically significant differences between treatment groups.

The primary endpoint of change from baseline to Week 52 in total score of the NSAA was chosen for its validated utility in measuring functional motor abilities in ambulatory children with DMD. By enrolling ambulatory patients, this study focused on pamrevlumab’s efficacy in DMD patients who had not yet experienced disease progression or musculoskeletal deterioration to the point of loss of ambulation. The minimum clinically important difference of the NSAA is estimated to be 2.3 to 2.9 points [23]. According to real-world and natural history studies, the NSAA typically decreases between 2.5 and 4 points per year after the age of 7 years for patients with DMD [24–26]. The changes observed in both treatment groups in this study were consistent with normal disease progression.

Secondary and exploratory endpoints were chosen both to support primary efficacy findings and to examine the effect of pamrevlumab on musculoskeletal and pulmonary function. None of the secondary endpoints nor the exploratory endpoints demonstrated statistically significant differences between treatment groups. The results of the subgroup analysis of patients with TTSTAND ≤5 seconds suggest that patients with less musculoskeletal deterioration and less disease progression may respond better to placebo than to pamrevlumab. However, the clinical significance of the results remains unclear due to the small sample size, and this finding contradicts that of the companion LELANTOS-1 study, which presented a subgroup of patients with less overall deterioration responding more favorably to pamrevlumab than placebo [22].

The incidence of TEAEs was high but balanced between the two treatment groups, which likely reflects the underlying symptoms of DMD and the AEs of long-term corticosteroid use. Overall, safety data were in line with expectations and did not reveal any new safety concerns. Pamrevlumab was generally safe and well tolerated.

This study had several limitations. First, analyses of the primary endpoint included patients with DMD that had not yet progressed to loss of ambulation. Further, the subgroup analyses contained small numbers of patients, which limited interpretation and generalization of the findings. In addition, longer periods of evaluation may be needed to establish significant differences, particularly for outcomes related to respiratory function. Finally, DMD progression has been attributed to genotype, glucocorticoid regimen, and other baseline and demographic characteristics [24, 27]. Because of the small sample sizes available for different genotypes in this study, it was not possible to determine if efficacy or safety varied according to genotype, which limited the interpretation of the findings.

## Acknowledgements

The authors would like to express their gratitude to the patients and their families for participating in this study. The authors would also like to thank Eugenio Mercuri for his participation and support during the trial. Medical writing support was provided by Jennifer L. Gibson, PharmD, of Kay Square Scientific (Newtown Square, PA, USA). This support was fully funded by FibroGen.

## Declaration of Funding

Funding for this study was provided by FibroGen, Inc. (San Francisco, CA, USA).

## Ethical Compliance

Informed consent was obtained from each patient or their legal guardian, and the study was approved by the respective Institutional Review Boards of each participating study site. This study was conducted in accordance with the Declaration of Helsinki, Good Clinical Practice (GCP), the International Council for Harmonisation (ICH) E6 Guidance for GCP, and any other applicable local health and regulatory requirements.

## Conflicts of Interest

**BLW, YP, YD, VAS, SW,** and **ZC** report nothing to disclose. **HCP** serves as a principal investigator for Applied Therapeutics, Avidity, Biogen, Biohaven, Dyne, Edgewise, Italfarmaco, NMD Pharma, NS Pharma, PepGen, Sarepta, Stealth, Takeda, Ultragenyx, and Zynerba. She also serves on the advisory board for Tyra and an advisory committee for the FDA. **SDL** has served as a PI for FibroGen and Genethon, on an advisory board for Solid, and as a consultant for Effik/Italfarmaco Group. **CC** served as a site investigator for AMO, Biogen, Dyne, Italfarmaco, Pfizer, Roche, PTC, Sarepta, and Wave, and a member of the DSMB for PepGen, Edgewise, and Solid. His hospital was a site for the clinical trial of this investigational therapy. **CC** is also an Editorial Board member of this journal, but was not involved in the peer-review process nor had access to any information regarding its peer review. **SD** has received personal fees for participating in advisory boards for UCB, Amikus, AstraZeneca, Argenx, Janssen Therapeutics, Amylyx, Biogen, Alexion, Alnylam, and CSL Behring, and providing expert testimony to Roche and Biogen. Fees were paid from FibroGen to her institution as part of the conduct of the LELANTOS-2 study, and she has received speaker honoraria from Zambon. **EC** is an employee of FibroGen and owns stock options. **OVG** was an employee of FibroGen at the time of the study, with restricted stock grants. **JFB** has received personal fees from AveXis/Novartis, Biogen, Dyne, Edgewise, FibroGen, Genentech, Momenta/Janssen, NS Pharma, Pfizer, PTC Therapeutics, Sarepta, Scholar Rock, Takeda, and WaVe. He has received grants from Alexion, Astellas, AveXis/Novartis, Biogen, Biohaven, Catabasis, CSL Behring, Cytokinetics, Dyne, Fibrogen, Genentech, ML Bio, Pfizer, PTC Therapeutics, Sarepta, and Scholar Rock.

## Data Availability Statement

FibroGen, Inc., is committed to data sharing and to furthering medical research and patient care. Based on scientific merit, requests from qualified external researchers for anonymized patient-level and study-level clinical trial data (including redacted clinical study reports) for medicines and indications approved in the United States and Europe will be considered after the respective primary study is accepted for publication. All data provided are anonymized to respect the privacy of patients who have participated in the trial in line with applicable laws and regulations.

## Notes

### Clinical Trial

NCT04632940

### Author Declarations

Informed consent was obtained from each patient or their legal guardian, and the study was approved by the respective Institutional Review Boards of each participating study site. This study was conducted in accordance with the Declaration of Helsinki, Good Clinical Practice (GCP), the International Council for Harmonisation (ICH) E6 Guidance for GCP, and any other applicable local health and regulatory requirements. See Table S1 in supplement for full details.

