## Supplemental Materials for "Pamrevlumab did not meet its primary endpoint for ambulatory patients with Duchenne Muscular Dystrophy: the LELANTOS-2 trial"

**LELANTOS-2: SUPPLEMENTARY MATERIAL**

**Methods**

Inclusion criteria:

I. Age and consent
   a. Males aged 6 to <12 years of age, ambulatory at screening initiation
   b. Written consent by patient and legal guardian as per regional/country and/or Institutional Review Board (IRB)/Independent Ethics Committee (IEC) requirements

II. DMD diagnosis: Medical history included diagnosis of DMD and confirmed Duchenne mutation using a validated genetic test

III. Pulmonary criteria
   a. Average (of screening and Day 0) percent predicted forced vital capacity (ppFVC) >45%
   b. On a stable dose of systemic corticosteroids for a minimum of 6 months, with no substantial change in dosage for a minimum of 3 months (except for adjustments for changes in body weight) prior to screening. Corticosteroid dosage had to be in compliance with the DMD Care Considerations Working Group recommendations (e.g., prednisone or prednisolone 0.75 mg/kg per day or deflazacort 0.9 mg/kg per day) or stable dose. A reasonable expectation was that dosage and dosing regimen would not change significantly for the duration of the study.

IV. Performance criteria

a. Able to complete 6-minute walking distance test (6MWD) with a distance of at least 270 meters but no more than 450 meters on two occasions within 3 months prior to randomization with ≤10% variation between these two tests
b. Able to rise (Time to Stand [TTSTAND]) from floor in <10 seconds (without aids/orthoses) at screening visit
c. Able to undergo magnetic resonance imaging test for the lower extremities vastus lateralis muscle

V. Vaccination: Agreement to receive annual influenza vaccinations during the study

VI. Laboratory criteria

a. Adequate renal function: cystatin C ≤1.4 mg/L
b. Adequate hematology and electrolytes parameters:
    i. Platelets >100,000/mcL
    ii. Hemoglobin >12 g/dL
    iii. Absolute neutrophil count >1500/μL
    iv. Serum calcium (Ca), potassium (K), sodium (Na), magnesium (Mg), and phosphorus (P) levels were within a clinically accepted range for DMD patients
c. Adequate hepatic function
    i. No history or evidence of liver disease
    ii. Gamma-glutamyl transferase (GGT) ≤3× upper limit of normal (ULN)
    iii. Total bilirubin ≤1.5× ULN

Exclusion criteria:

I. General criteria

a. Concurrent illness other than DMD that could cause muscle weakness and/or impairment of motor function

b. Severe intellectual impairment (e.g., severe autism, severe cognitive impairment, severe behavioral disturbances) preventing the ability to perform study assessments in the investigator's judgment

c. Previous exposure to pamrevlumab

d. BMI ≥40 kg/m2 or weight >117 kg

e. History of

   i. Allergic or anaphylactic reaction to human, humanized, chimeric, or murine monoclonal antibodies

   ii. Hypersensitivity to study drug or any component of study drug

f. Exposure to any investigational drug (for DMD or not), in the 30 days prior to screening initiation or use of approved DMD therapies (e.g., eteplirsen [exondys 51], ataluren, golodirsen [vyondys 53], casimersen [amondys 45]) within 5 half-lives of screening, whichever was longer, with the exception of the systemic corticosteroids, including deflazacort

II. Pulmonary, renal, and cardiac criteria

a. Required ≥16 hours continuous ventilation

b. Poorly controlled asthma or underlying lung disease such as bronchitis, bronchiectasis, emphysema, or recurrent pneumonia that, in the opinion of the investigator, could have impacted respiratory function

c. Hospitalization due to respiratory failure within the 8 weeks prior to screening

d. Severe uncontrolled heart failure (NYHA Classes III-IV) or renal dysfunction, including any of the following:
   i. Need for intravenous diuretics or inotropic support within 8 weeks prior to screening
   ii. Hospitalization for a heart failure exacerbation or arrhythmia within 8 weeks prior to screening
   iii. Patients with glomerular filtration rate (GFR) of <30 mL/min/1.73m2 or with other evidence of acute kidney injury as determined by investigator
e. Arrhythmia requiring antiarrhythmic therapy
f. Any other evidence of clinically significant structural or functional heart abnormality
g. Clinical judgment: The investigator judged that the subject would be unable to fully participate in the study and complete the study for any reason, including inability to comply with study procedures and treatment, or any other relevant medical, surgical, or psychiatric conditions

**Table S1. Institutional review board and ethics approvals for LELANTOS-2**

| Sponsor | Protocol # | Country Name | Recipient Type | Recipient Name | Address | Site: Site # | Ethics approval? (granted/waived) |
|---|---|---|---|---|---|---|---|
| FibroGen | FGCL-3019-094 | AUSTRALIA | Central EC | Research Ethics & Governance RCH Human Research Ethics Committee The Royal Children's Hospital Melbourne | Level 4, South Building 50 Flemington Road Parkville Victoria 3052 Australia | N/A | Granted |
| FibroGen | FGCL-3019-094 | AUSTRIA | Central EC | Ethikkommission der Stadt Wien | 3 Thomas-Klestil-Platz 8 1030 Wien Austria | N/A | Granted |
| FibroGen | FGCL-3019-094 | BELGIUM | Local IEC | Commissie voor Medische Ethiek Universiteit Gent/UZ Gent | Commissie voor Medische Ethiek Universiteit Gent/UZ Gent C. Heymanslaan 10 9000 Gent | 8301 | Granted |
| FibroGen | FGCL-3019-094 | BELGIUM | Local IEC | Centre Hospitalier Régional de la Citadelle | Centre Hospitalier Régional de la Citadelle Boulevard du 12ème de Ligne 1 4000 Liège | 8303 | Granted |
| FibroGen | FGCL-3019-094 | BELGIUM | Central EC | Ethische Commissie Onderzoek UZ/KU Leuven UZ Leuven | Campus Gasthuisberg Herestraat 49 3000 Leuven Belgium | N/A | Granted |
| FibroGen | FGCL-3019-094 | CANADA | Local IRB/IEC | London Health Sciences Centre | 800 Commissioners Rd E, London, ON N6A 5W9, Canada | 8204 | Granted |

| | | | | | | | |
|---|---|---|---|---|---|---|---|
| FibroGen | FGCL-3019-094 | China | Site IRB/IEC | Drug Clinical Trial Ethics Committee of Peking Union Medical College Hospital, Chinese Academy of Medical Sciences | No.41, Dacang Hutong, Xicheng District, Beijing City,China | 8601 | Granted |
| FibroGen | FGCL-3019-094 | China | Site IRB/IEC | Clinical Trial Ethics Committee of West China Second University Hospital, Sichuan University | No. 20, Section 3, Renmin South Road, Chengdu City, Sichuan Province,China | 8603 | Granted |
| FibroGen | FGCL-3019-094 | China | Site IRB/IEC | Institutional Review Board of Children's Hospital of Chongqing Medical University | No.136, Zhongshang 2nd road, Yuzhong District, Chongqing City, China | 8604 | Granted |
| FibroGen | FGCL-3019-094 | China | Site IRB/IEC | Clinical Drug, Device and New medical technologies Ethics Committee of the 1st Affiliated Hospital, Sun Yat- sen University | Room 110, 1 floor, Longzhu Buliding, No.5, Zhusigangerma Road, Guangzhou City,Guangdong Province, China | 8605 | Granted |
| FibroGen | FGCL-3019-094 | China | Site IRB/IEC | Medical Ethics Committee of Xiangya Hospital Central South University | No.87, Xiangya Road, Changsha City, Hunan Province, China | 8606 | Granted |
| FibroGen | FGCL-3019-094 | FRANCE | Central EC | Comité de Protection des Personnes - Nord Ouest II | Bâtiment Pharmacie - Hôpital Nord, Place Victor Pauchet, 80054 Amiens, France | N/A | Granted |
| FibroGen | FGCL-3019-094 | ITALY | Local IEC | Comitato Etico IRCCS Ospedale San Raffaele | Via Olgettina 60, 20132, Milano (MI), Italy | 8222 | Granted |
| FibroGen | FGCL-3019-094 | ITALY | Local IEC | Comitato Etico Milano Area 3 | Piazzale Ospedale Maggiore, 3, 20162, Milano (MI), Italy | 8223 | Granted |

| | | | | | | | |
|---|---|---|---|---|---|---|---|
| FibroGen | FGCL-3019-094 | ITALY | Local IEC | Comitato Etico dell'IRCCS Ospedale Pedi-atrico Bambino Gesù | Viale di Villa Pamphili 84-100, 00155, Roma (RM), Italy | 8224 | Granted |
| FibroGen | FGCL-3019-094 | ITALY | Local IEC | Comitato Etico IRCCS E.Medea - Sez.Scientifica Associazione La Nostra Famiglia | Via Don Luigi Monza, 20, 23842, Bosisio Parini (LC), Italy | 8225 | Granted |
| FibroGen | FGCL-3019-094 | ITALY | Central EC | Comitato Etico Territoriale (CET) - Comitato Etico Lazio Area 3 c/o Segreteria Tecnico Scientifica all’Ex Collegio Ioanneum | primo piano stanza 221 Largo F. Vito 1<br>00168 Roma Italy | N/A | Granted |
| FibroGen | FGCL-3019-094 | NETHERLANDS | Central EC | Commissie Mensgebonden Onderzoek regio Arnhem-Nijmegen | p/a Radboudumc, Huispost 628, PO box 9101,<br>6500 HB Nijmegen The Netherlands | N/A | Granted |
| FibroGen | FGCL-3019-094 | SPAIN | Central EC | Hospital Clínico San Carlos Comité de Ética<br>de la Investigación con Medicamentos | C/Profesor Martín Lagos, s/n. - Puerta G -<br>4ª Norte Madrid 28040 Madrid Spain | N/A | Granted |
| FibroGen | FGCL-3019-094 | UNITED KINGDOM | Central EC | South West – Central Bristol Research Ethics Committee (REC) | Ground Floor Temple Quay House, 2 The Square<br>Bristol, BS1 6PN, United Kingdom | N/A | Granted |
| FibroGen | FGCL-3019-094 | UNITED STATES | Local IRB/IEC | Cincinnati Children’s Institutional Review Board | 3333 Burnet Avenue<br>Cincinnati, Ohio, 45229 | 7901 | Granted |

| | | | | | | | |
|---|---|---|---|---|---|---|---|
| FibroGen | FGCL-3019-094 | UNITED STATES | Local IRB/IEC | Cincinnati Children's Hospital Medical Center | 3333 Burnet Avenue Cincinnati, Ohio, 45229 | 7901 | Granted |
| FibroGen | FGCL-3019-094 | UNITED STATES | Local IRB/IEC | Children's Hospital of Philadelphia | 34th Street and Civic Center Boulevard Philadelphia, Pennsylvania, 19104 | 7908 | Granted |
| FibroGen | FGCL-3019-094 | UNITED STATES | Local IRB/IEC | Children's Hospital of Philadelphia Institutional Review Board | 3401 Civic Center Boulevard Philadelphia, Pennsylvania, 19104 | 7908 | Granted |
| FibroGen | FGCL-3019-094 | UNITED STATES | Local IRB/IEC | John Hopkins Medicine - Office of Human Subjects Research | 1620 McElderry Street East Baltimore Campus Baltimore, Marlyand, 21205-1911 | 7959 | Granted |
| FibroGen | FGCL-3019-094 | UNITED STATES | Local IRB/IEC | Kennedy Krieger Institute | 707 North Broadway Baltimore, Maryland, 21205 | 7959 | Granted |
| FibroGen | FGCL-3019-094 | UNITED STATES | Local IRB/IEC | University of Utah Institutional Review Board | The Research Administration Building 75 South 2000 East Salt Lake City, Utah 84112 | 7962 | Granted |
| FibroGen | FGCL-3019-094 | UNITED STATES | Local IRB/IEC | Vanderbilt University Medical Center | Human Research Protections Program 3319 West End Ave Suite 600 Nashville, TN 37203 | 7968 | Granted |
| FibroGen | FGCL-3019-094 | UNITED STATES | Local IRB/IEC | University of California San Diego Human Research Protections Program | 9500 Gilman Drive, La Jolla, CA 92093- 0021 | 7975 | Granted |
| FibroGen | FGCL-3019-094 | UNITED STATES | Central IRB/IEC | Advarra | 6940 Columbia Gateway Drive Columbia, Maryland, 21046 | N/A | Granted |

| | | | | | | | |
|---|---|---|---|---|---|---|---|
| FibroGen | FGCL-3019-094 | UNITED STATES | Central IRB/IEC | WCG IRB | 1019 39th Ave SE Suite 120 Puyallup, WA 98374 | N/A | Granted |
| FibroGen | FGCL-3019-094 | UNITED STATES | Central IRB/IEC | WCG IRB - Copernicus Group Independent Review Board | 1019 39th Ave SE Suite 120 Puyallup, WA 98374 | N/A | Granted |
